# Ambient Only vs. Longitudinal Data-Enhanced AI Documentation: A Pilot Study Quantifying the Value of Historical Clinical Context in Primary Care

**DOI:** 10.1101/2025.11.07.25339620

**Authors:** Michael Zuckerman, Gal Eyal, Roei Magen, Nir Lewis, Omer Harnof, Shiri Shifman, Zach Avraham, Lynn Joffe, Kevin Gallagher, Yair E. Lewis

## Abstract

**Background:** Ambient artificial intelligence (AI) clinical documentation tools have gained rapid adoption in healthcare to address physician burnout from documentation burden. However, current implementations primarily rely on real-time audio capture without systematically incorporating longitudinal patient data, potentially limiting documentation completeness for chronic disease management.

**Objective:** To compare documentation completeness between ambient audio-only workflows and those augmented with historical clinical data from electronic health records (EHRs) for type 2 diabetes and hypertension encounters in primary care.

**Methods:** We conducted a retrospective, paired, cross-sectional study of 354 primary care encounters in which diabetes mellitus (DM, n=119) and/or hypertension (HTN, n=281) were treated. Each condition instance was analysed twice to compare two methods of automated documentation: using only physician-patient conversation transcripts (termed “ambient only”) compared with consolidated automated documentation that includes historical clinical data in addition to the ambient conversation (ambient + history; termed “consolidated”). Documentation completeness was assessed using the “assessment” subset of the QNOTE clinical documentation quality measurement instrument, evaluating four domains: completeness, clinical coherence, clarity, conciseness. Scoring was automated using an LLM pipeline with physician validation on a 20% sample.

**Results:** Consolidated documentation achieved significantly higher mean total assessment composite score compared to ambient-only (94.8 vs. 80.1 on a scale of 0-100; difference 14.6 points; 95% CI 13.4-15.8; P<0.001). The largest improvements was observed in the completeness domain (difference 42.5 points; P<0.001). DM and HTN both showed similar performance of consolidated documentation vs. ambient only.

**Conclusions:** Augmenting ambient AI documentation with historical EHR data significantly improves documentation completeness for chronic disease management in primary care. These preliminary findings challenge the prevailing audio-first implementation paradigm and suggest that bidirectional EHR integration may be essential for comprehensive AI-assisted documentation, particularly for conditions requiring synthesis of longitudinal clinical data.

## Background

Clinical documentation has emerged as a primary driver of physician burnout and dissatisfaction in modern healthcare. Studies have demonstrated that physicians spend a substantial amount of their workday engaging with electronic health records (EHRs) [1], with documentation consuming up to two hours for every hour of direct patient care[2]. This burden has been one of the drivers of the rapid adoption of ambient artificial intelligence (AI) clinical documentation tools, with rapid adoption across the US healthcare system[3,4].

Ambient clinical documentation leverages natural language processing and generative AI to passively capture patient-clinician conversations and automatically generate structured clinical notes. Healthcare organisations implementing these solutions report multiple benefits including decreased physician burnout, improved same-day closure rates, and enhanced clinical workflows. The evidence base for time savings is growing but heterogeneous. A University of Pennsylvania study found a 20.4% reduction in time spent per appointment in notes (from 10.3 to 8.2 minutes)[5], while similar implementations at Sutter Health demonstrated a 14.5% decrease (from 6.2 to 5.3 minutes)[6]. Critically, these tools reduce after-hours documentation by 2.5 to 3 hours weekly, directly addressing “pajama time” that erodes work-life balance[5].

Beyond time metrics, ambient scribing impacts documentation quality and completeness, though results vary considerably. Early evaluations suggest that AI-generated notes capture more detailed histories of present illness and may include clinical information that physicians might otherwise omit for brevity[7]. However, ambient scribing systems face fundamental limitations in capturing the full clinical encounter; they cannot document physical examination findings unless explicitly verbalised by the clinician, miss non-verbal cues such as patient affect or subtle physical signs, and fail to capture clinical observations that physicians naturally observe but may not articulate during the visit. This reliance on spoken content alone creates systematic gaps in documentation that are particularly problematic for comprehensive chronic disease management. The requirement for diligent clinician oversight introduces “editing fatigue” and automation bias risks, potentially undermining efficiency gains[8].

This reliance on audio-only workflows represents a critical gap in chronic disease management, where comprehensive documentation requires synthesis of current observations with historical trends, laboratory values, and treatment responses. For conditions like diabetes and hypertension, quality metrics demand documentation of glycemic control trajectories, medication adjustments, screening for complications, and preventive care measures-elements that may not surface naturally in routine conversations. In addition, incomplete documentation can impact provider compensation. The Medicare Evaluation and Management (E/M) documentation requirements, particularly the MEAT (Monitoring, Evaluation, Assessment, Treatment) criteria for chronic conditions, necessitate explicit linkage between current findings and historical data to support medical necessity and care continuity.

Recent technical developments suggest that integrating historical context is feasible, with studies demonstrating that large language models can effectively synthesise multiple data sources for clinical documentation. However, systematic evaluation of how historical data augmentation affects documentation completeness, particularly for chronic disease management, remains unexplored in the peer-reviewed literature.

This study addresses this gap by directly comparing documentation completeness between ambient audio-only workflows and those augmented with historical clinical data from EHRs and health information exchanges (HIE). By utilising a subset of the QNOTE clinical documentation quality scoring framework [9], and applying it to diabetes and hypertension encounters, we provide the first empirical assessment of whether incorporating longitudinal patient context enhances the quality of AI-generated clinical documentation. This evaluation is particularly timely given mixed evidence on time savings and growing recognition that persistent chart review needs may offset efficiency gains when critical historical information is omitted from ambient-only documentation.

## Methods

### Study Design

We conducted a retrospective, paired, cross-sectional study to evaluate the completeness of primary care documentation generated through two methods. Each clinical encounter was documented twice: once using only the physician–patient conversation transcript (ambient-only), and once augmented with structured data from the electronic health record (ambient+history; “consolidated”). We compared the two approaches using a subset of the validated QNOTE scoring framework [9] (Figure 1).

**Figure 1.**
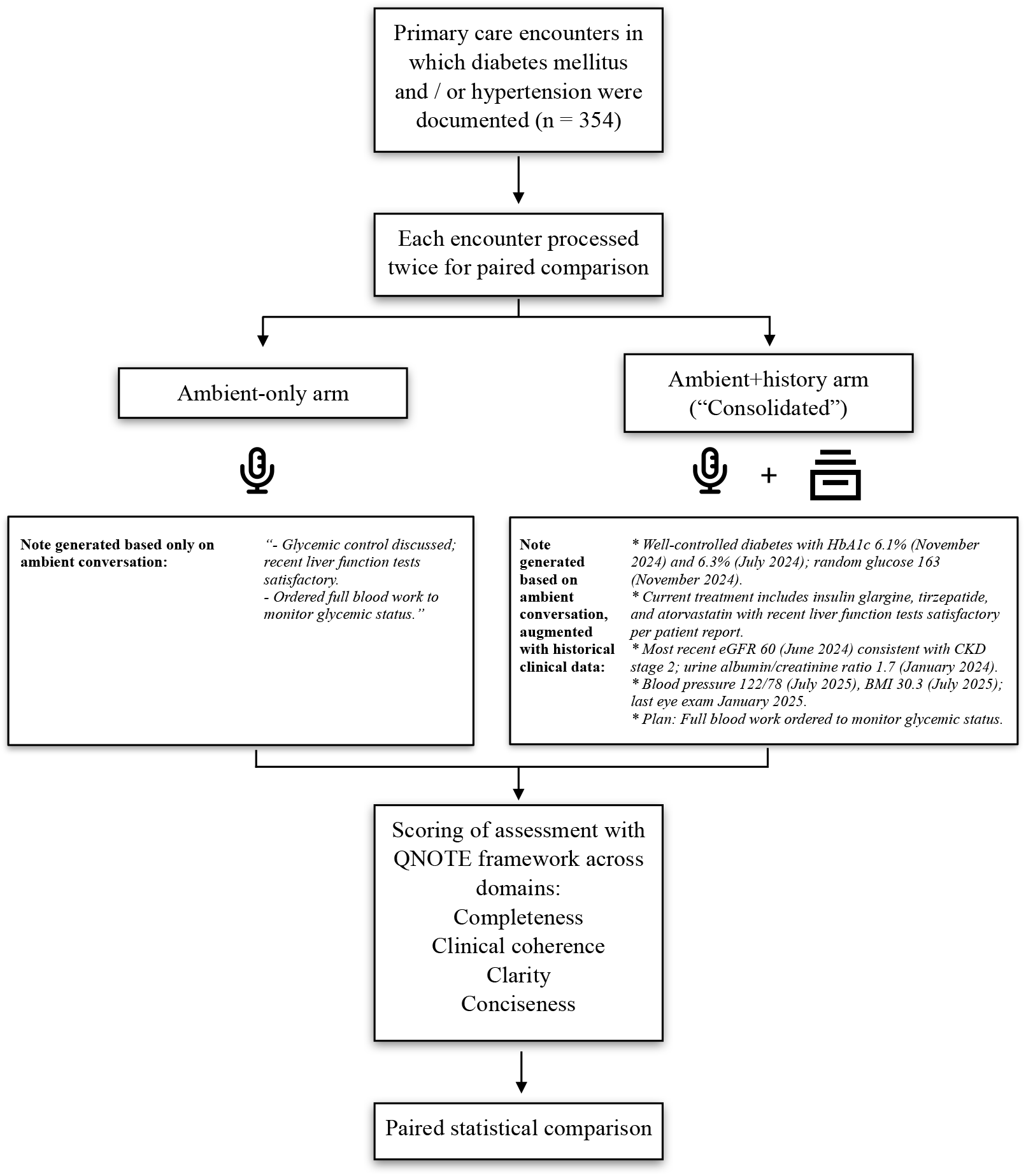
Study Design

### Setting and Participants

The study included de-identified AI generated ambient clinical scribing notes of primary care encounters from two primary care clinics in the United States. We included encounters in which for type 2 diabetes mellitus and hypertension were treated, selecting based on the ICD-10 codes that were coded in the encounter. Only the “assessment and plan” section of the encounter was evaluated, as per study goal. All encounters meeting the pre-specified inclusion criteria during the study period were included; no sampling or additional selection was undertaken. Although visits often addressed multiple conditions, we analysed only documentation elements related to those two conditions mentioned above, as a proxy for chronic disease documentation.

### Documentation Generation

Ambient-only documentation was generated by an ambient clinical documentation platform (Navina, NY, USA) that produces structured SOAP notes directly from the ambient patient-physician interaction. As mentioned above, only the assessment and plan section was used in this study. Consolidated documentation was created by an combining the ambient documentation with longitudinal patient data from the electronic health record (EHR), including medications, laboratory values, family history, and consultation notes. The input was processed through an automated retrieval-augmented generation (RAG) pipeline which integrated the conversational and historical clinical data into a unified clinical note. The selection of relevant clinical data elements for each condition was based on a knowledge graph manually curated by physician domain experts, which enumerated condition-to-evidence linkages (e.g., key labs, medications, problem lists, imaging, and consult notes) used by the retriever to filter and rank candidate facts. The foundation model used in creating the consolidated note was Claude Sonnet 4 (Anthropic, CA, USA).

### Scoring Framework

Documentation quality was assessed using a modified version of the QNOTE instrument, a validated tool for evaluating the quality of clinical notes in electronic health records [9]. Specifically, we adapted Section 10 (Assessment) of the QNOTE framework, which focuses on the quality of assessment section of the encounter documentation, across four domains (Table 1). We operationalised this section into 14 sub-questions. Each sub question in the instrument was scored according to the following scale: “Full” (100 points); “Partial” (50 points); or “Unacceptable” (0 points). Domain scores were calculated as the arithmetic mean of all items within each respective domain, regardless of the number of items per domain. The overall composite score was derived using an equal-weighting approach across all four domains. Specifically, each domain contributed 25% to the final composite score, calculated as the unweighted mean of the four domain scores.

**Table 1.** QNOTE Instrument - Assessment Section.

| **Question Number** | **Domain** | **Sub-Question** |
| --- | --- | --- |
| 1 | Completeness | Does the assessment include an explicit statement about condition status (e.g., controlled/stable/not at goal/worsening)? |
| 2 | Completeness | Does it mention a measurement of the core disease marker (e.g., HbA1c for diabetes, BP for hypertension, EF for heart failure)? |
| 3 | Completeness | Does it mention the medications the patient is taking for this condition? |
| 4 | Completeness | Does it mention changes in the medication plan OR a rationale for not changing the current medication plan? |
| 5 | Completeness | Does it mention relevant vital sign measurements (BP, HR, weight/BMI, SpO2)? |
| 6 | Completeness | Does it mention relevant past medical history for this condition? |
| 7 | Completeness | Does it mention condition-relevant complication monitoring (e.g., kidney function, lipid panel, eye/foot exam for diabetes)? |
| 8 | Completeness | Does it mention condition-relevant comorbidities or contextual factors? |
| 9 | Clinical coherence | Clinical reasoning logical |
| 10 | Clinical coherence | Free of contradictions or errors |
| 11 | Clarity | Language clear and unambiguous |
| 12 | Clarity | Information organised clearly and logically |
| 13 | Clarity | Easily understandable to other providers |
| 14 | Conciseness | Is the note concise, readable, and free of redundant information |

Full credit is achievable even when elements are not applicable (e.g., documenting “no complications” or “not taking any medications”). Scoring was done via a Python pipeline, utilising GPT-5 (OpenAI, CA, USA) to score the modified QNOTE framework as described above.

### Scoring Validation

To validate scoring reliability, two medical subject-matter experts rated the same random sample of notes (n = 20) using the modified QNOTE instrument described above. Both experts scored each note independently and in a blinded fashion, without knowledge of whether the note originated from the ambient-only or consolidated version. Agreement was assessed both between human raters (inter-reviewer agreement) and between the human consensus score and the LLM-generated score (model– human agreement), to confirm the validity and reproducibility of the automated evaluation. We calculated agreement at two thresholds: exact agreement (identical scores) and substantial agreement (score differences <50 points). Inter-rater exact agreement was 78.4% and substantial agreement was 95.3%. Human-model exact agreement was 73.8% and substantial agreement was 91.9%.

### Statistical Analysis

All statistical analyses were performed using R (version 4.3.2; R Core Team, 2023). Given our paired cross-sectional design where each condition instance was analysed using both documentation methods, we employed paired Student’s t-tests to compare QNOTE assessment scores between ambient-only and consolidated (ambient + history) documentation.

Descriptive statistics were calculated for both documentation methods across all four QNOTE assessment domains (completeness, clinical coherence, clarity, and conciseness) as well as the total assessment composite score. For each domain score and the composite score, 95% confidence intervals were computed for mean estimates within each documentation arm. Analyses were stratified by condition and conducted at three levels: (1) diabetes mellitus encounters (n=119), (2) hypertension encounters (n=281), and (3) the combined dataset. Given that some encounters included treatment for both conditions, each condition instance was analysed independently according to our paired design.

To account for multiple comparisons across the four domains and composite score within each analysis stratum, we applied the Benjamini-Hochberg false discovery rate (FDR) correction. The statistical significance threshold was set at p < 0.05 (two-tailed).

## Results

### Overall score

We evaluated 354 primary care encounters documenting management of type 2 diabetes mellitus (DM, n=119 condition instances) and essential hypertension (HTN, n=281 condition instances). Some encounters addressed both conditions. Each condition instance was independently documented using both methods (ambient-only and consolidated), creating paired comparisons for analysis.

The overall mean assessment score was substantially higher for consolidated documentation than for ambient-only documentation (Table 2 and Figure 2; 94.8 vs. 80.1; difference, 14.6 points; 95% CI,13.4 to 15.8; adjusted P<0.001). Statistically significant improvements were observed across all domains. The most significant gains occurred in completeness (difference, 42.5 points; adjusted P<0.001). Results were consistent across analysis of both conditions, DM and HTN (Table 2).

**Table 2.** Results: Overall Assessment Score.

| Pairs |  | Scores |  | 95% CI | p<sub>adj</sub> |
| --- | --- | --- | --- | --- | --- |
|  |  | Ambient | Consolidated | Δ (Consolidated – Ambient) |  |
| All |  |  |  |  |  |
| Final Assessment Score (Composite) | 400 | 80.1 | 94.8 | 14.6 [13.4, 15.8] | <0.001 *** |
| Completeness | 400 | 40.4 | 82.9 | 42.5 [40.5, 44.6] | <0.001 *** |
| Clinical Coherence | 400 | 91.6 | 98.9 | 7.3 [6.0, 8.7] | <0.001 *** |
| Clarity | 400 | 93.9 | 99.3 | 5.5 [3.9, 7.1] | <0.001 *** |
| Conciseness | 400 | 96.2 | 99.4 | 3.1 [1.7, 4.5] | <0.001 *** |
| DM |  |  |  |  |  |
| Final Assessment Score (Composite) | 119 | 81.3 | 96.0 | 14.7 [12.5, 16.8] | <0.001 *** |
| Completeness | 119 | 44.5 | 86.4 | 41.8 [38.2, 45.5] | <0.001 *** |
| Clinical Coherence | 119 | 93.3 | 99.6 | 6.3 [4.2, 8.4] | <0.001 *** |
| Clarity | 119 | 93.9 | 99.7 | 5.8 [3.0, 8.5] | <0.001 *** |
| Conciseness | 119 | 95.0 | 99.6 | 4.6 [2.0, 7.3] | <0.001 *** |
| HTN |  |  |  |  |  |
| Final Assessment Score (Composite) | 281 | 79.6 | 94.3 | 14.6 [13.2, 16.1] | <0.001 *** |
| Completeness | 281 | 38.6 | 81.4 | 42.8 [40.4, 45.3] | <0.001 *** |
| Clinical Coherence | 281 | 90.9 | 98.7 | 7.7 [6.0, 9.4] | <0.001 *** |
| Clarity | 281 | 93.8 | 99.2 | 5.4 [3.4, 7.3] | <0.001 *** |
| Conciseness | 281 | 96.8 | 99.3 | 2.5 [0.8, 4.2] | <0.01 ** |

**Table 3.**
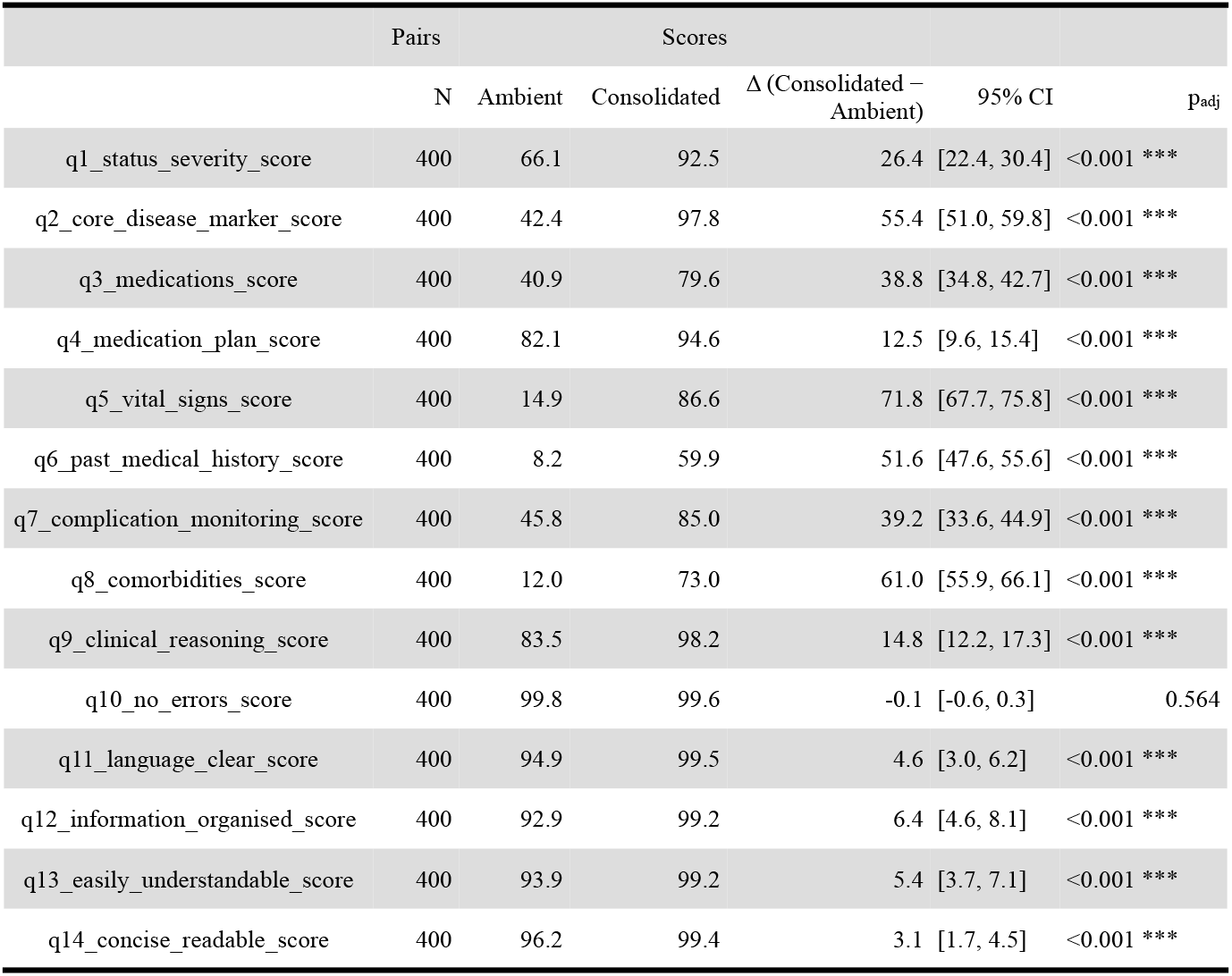
Results: Sub-Domain Level.

**Figure 2.**
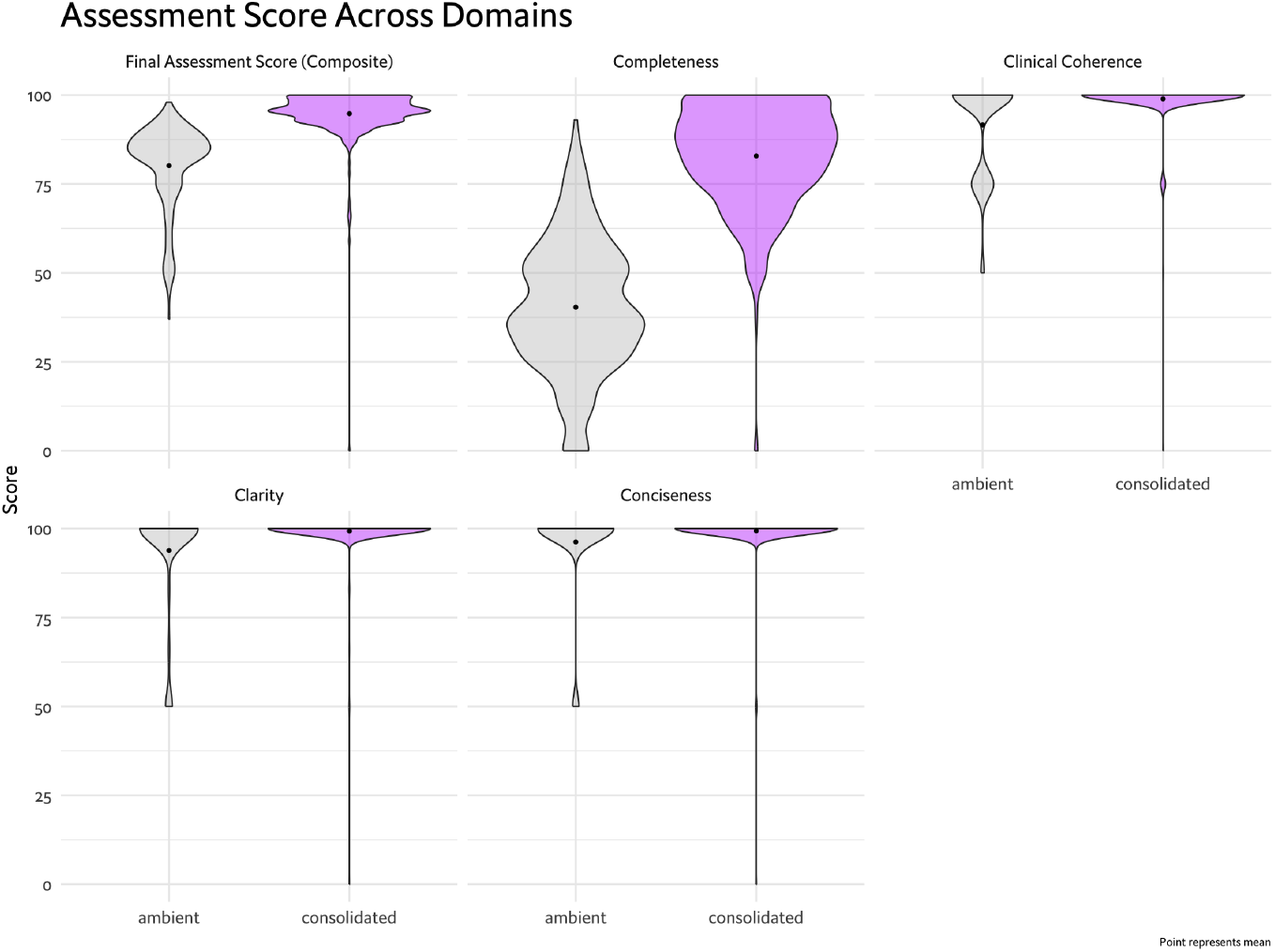
Results - Overall Assessment Score

Nearly all encounter pairs showed higher scores with historical clinical data augmentation, indicating a consistent benefit of multimodal documentation.

### Subgroup analysis at the sub-domain level

We analysed the results at the sub-domain level as well (Table 3 and Figure 3). The largest differences were observed for vital signs (q5; 86.6 vs. 14.9; difference, 86.6 points), comorbidities (q8; 73.0 vs. 12.0; difference, 61.0 points), and past medical history (q6; 59.9 vs. 8.2; difference, 51.6 points). Core disease markers (q2; 97.8 vs. 42.4; difference, 55.4 points) and disease severity (q1; 100.0 vs. 56.8; difference, 43.2 points) also demonstrated marked improvements. Of note, there was marked variability in documentation quality for ambient-only, as opposed to much narrower distribution in the consolidated note group. This likely reflects variance in amount of conversation between encounters.

**Figure 3.**
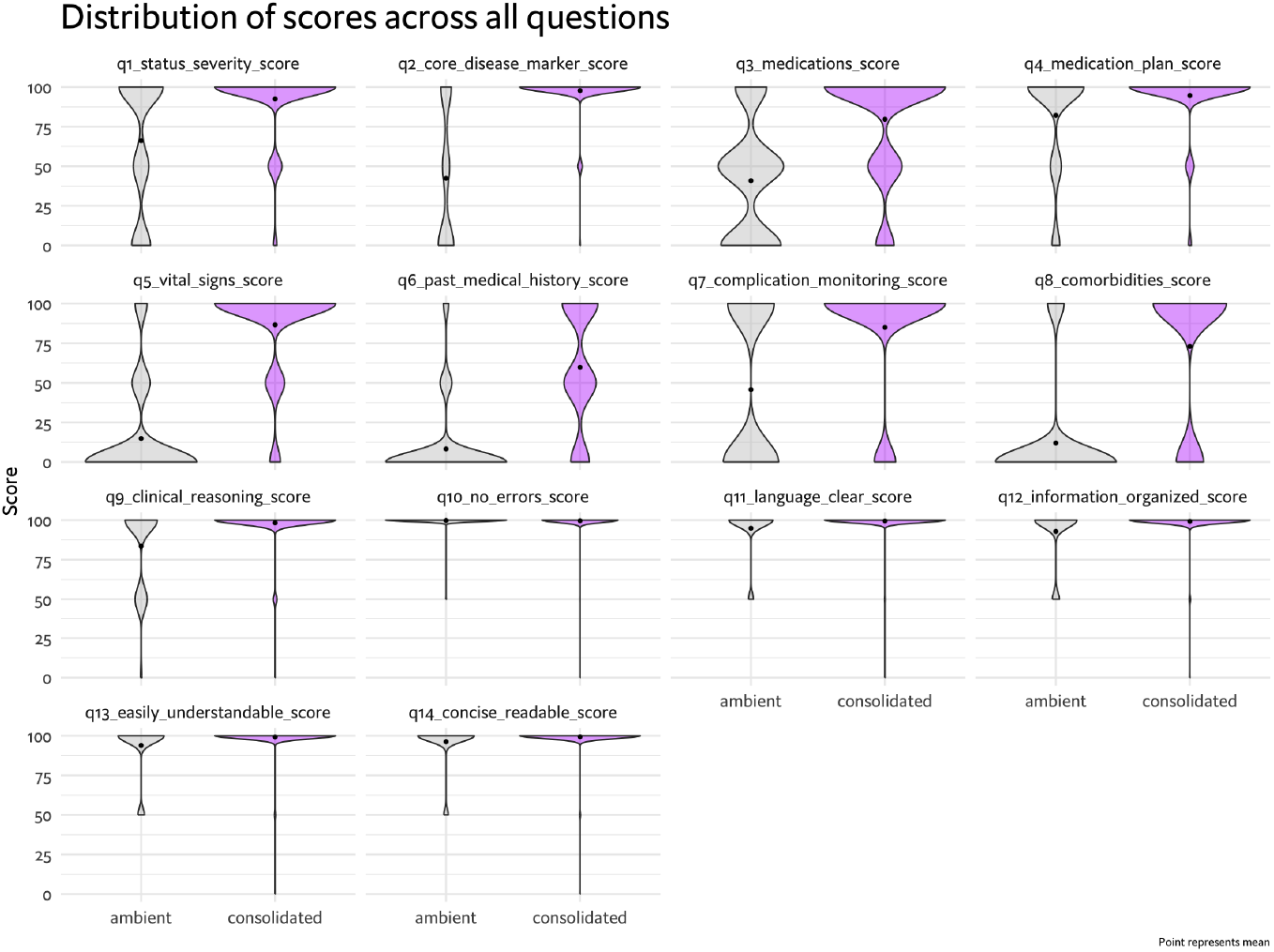
Results - Sub-Domain Level

## Discussion

This pilot study provides initial evidence suggesting that augmenting ambient AI documentation with historical clinical data may improve documentation completeness for chronic disease management in primary care. The 14.6-point improvement in overall assessment score represents a potentially meaningful enhancement, particularly given the 42.5-point gain in the “completeness” domain - a critical aspect of quality of clinical documentation.

Our findings relate to a fundamental limitation of audio-only ambient documentation: the verbal patient-physician interaction may omit quantitative and historical information essential for comprehensive chronic care. The substantial differences between ambient-only and consolidated notes in the completeness domains of core disease markers, vital signs, and condition relevant comorbidities demonstrate that critical elements like laboratory trends, vital sign patterns, and complication screening results rarely surface naturally in clinical conversations.

The minimal impact on medication plan and clinical reasoning documentation suggests that treatment discussions are already well-captured through conversation alone. This finding aligns with workflow observations that medication changes and care planning constitute major components of verbal physician-patient interactions.

These preliminary results have important implications for ambient AI implementation strategies. While current deployments emphasize real-time transcription efficiency, our data suggest that organisations should prioritise bidirectional EHR integration, i.e. the ability to retrieve historical clinical data, to realise the full potential of AI documentation assistance.

The study also illuminates the tension between documentation efficiency and completeness. Although ambient-only tools demonstrably reduce documentation time, the resulting notes may require substantial post-encounter chart review and editing to meet quality and reimbursement standards - potentially offsetting some of the initial time savings. Historical data integration could improve efficiency by reducing the need for manual note augmentation, while ensuring compliance for E/M billing.

### Limitations

Several limitations warrant consideration. First, this pilot analysed a small dataset of encounters by a limited number of physicians, in two clinical sites, limiting generalisability. Second, we evaluated completeness rather than accuracy or clinical relevance. Finally, our analysis focused exclusively on diabetes and hypertension; conditions with different documentation requirements may show varying benefit patterns.

### Future Directions

This pilot study justifies more rigorous investigation into optimal AI documentation strategies. Future research using a larger and more diverse dataset should evaluate real-world outcomes including physician time savings when accounting for note review and editing, clinical decision-making impact, billing accuracy, and patient safety considerations. Investigating optimal strategies for presenting historical context - balancing comprehensiveness with cognitive load - will be crucial for successful implementation.

As ambient AI tools rapidly proliferate across healthcare systems, these preliminary findings underscore the importance of moving beyond simple transcription and toward intelligent documentation systems that synthesise conversational and historical, longitudinal, clinical data. While confirmatory studies are needed, this initial evidence suggests that integrating historical context may support more comprehensive documentation for chronic disease management in primary care.

## Data Availability

Data produced in the present study are available upon reasonable request to the authors, other than the de-identified clinical notes which cannot be shared in order to protect patient privacy and in accordance with HIPAA regulations.

## Ethics Statement

All data used in the study was fully de-identified prior to analysis. Study was determined as non-human subject research (NSHR) by ethics committees of both clinical sites (DTC, HHHN).

---

Table 3. Results: Sub-Domain Level

Figure 3: Results: Sub-Domain Level

